# Effects of Covid-19 on the human central olfactory system: a natural pre-post experiment

**DOI:** 10.1101/2021.12.27.21268455

**Authors:** Evelina Thunell, Moa G. Peter, Vincent Lenoir, Patrik Andersson, Basile N. Landis, Minerva Becker, Johan N. Lundström

## Abstract

Reduced olfactory function is the symptom with the highest prevalence in COVID-19 with nearly 70% of individuals with COVID-19 experiencing partial or total loss of their sense of smell at some point during the disease. The exact cause is not known but beyond peripheral damage, studies have demonstrated insults to both the olfactory bulb and central olfactory brain areas. However, these studies often lack both baseline pre-COVID-19 assessments and a control group and could therefore simply reflect preexisting risk factors. Right before the COVID-19 outbreak, we completed an olfactory focused study including structural MR brain images and a full clinical olfactory test. Opportunistically, we invited participants back one year later, including 9 participants who had experienced mild to medium COVID-19 (C19+) and 12 that had not (C19-), thereby creating a pre-post controlled natural experiment with a control group. Despite C19+ participants reporting subjective olfactory dysfunction, few showed signs of objectively altered function one year later. Critically, all but one individual in the C19+ group had reduced olfactory bulb volume with an average volume reduction of 14.3%, but this did not amount to a significant between group difference compared to the control group (2.3% reduction) using inference statistics. No morphological differences in cerebral olfactory areas were found but we found stronger functional connectivity between olfactory brain areas in the C19+ croup at the post measure. Taken together, these data suggest that COVID-19 might cause a long-term reduction in olfactory bulb volume but with no discernible differences in cerebral olfactory regions.

## INTRODUCTION

Olfactory dysfunction is a key symptom of COVID-19 (Gerkin et al., 2021). The reported prevalence of complete olfactory loss (anosmia) is about 50% with an additional 10-20% reporting less severe olactory dysfunction (Hannum et al., 2020; Iravani et al., 2020). Thus, nearly 70% of all individuals with COVID-19 experience partial or total loss of their sense of smell at some point during the disease. In accordance, complaints about a reduced sense of smell is the symptom with the highest odds ratio in non-hospitalized cases (Menni et al., 2020; Rudberg et al., 2020). Despite the clear clinical link between olfactory dysfunction and COVID-19, our understanding of the mediating mechanisms is, however, limited.

The main entry receptor for SARS-CoV-2, the virus that causes COVID-19, is thought to be the angiotensin converting enzyme 2 (ACE2) receptor which is expressed throughout the human respiratory system with high density in the nasal epithelium and especially in the supporting sustentacular cells (Fodoulian et al., 2020; Hou et al., 2020; Muus et al., 2021). Much like the SARS-CoV-1 and influenza viruses (Durrant, Ghosh, & Klein, 2016), SARS-CoV-2 can invade the central nervous system through the olfactory mucosa via a retrograde route (de Melo et al., 2021). SARS-CoV-2 nucleoproteins and associated inflammation have been detected in infected animal models along the entire olfactory route, from the olfactory sensory neurons to the olfactory bulb (OB; de Melo et al., 2021). In humans, there is indirect and mixed evidence of SARS-CoV-2 as a neurotropic virus. Studies have demonstrated post mortem brain pathologies after COVID-19 but without clear evidence of SARS-CoV-2 RNA presence (Mukerji & Solomon, 2021).

There is further conflicting or weak evidence of neuroinvasion within the olfactory system in humans with a dominance of case studies or assessment of severe cases. A post-mortem case study found low levels of virus RNA in the OB (Meinhardt et al., 2021), and several neuroimaging studies have demonstrated anosmia-related edemas and abnormalities evident in CT or MR images of the OB post-COVID-19 infection (Aragão, Leal, Cartaxo Filho, Fonseca, & Valença, 2020; Galougahi, Ghorbani, Bakhshayeshkaram, Naeini, & Haseli, 2020; Kandemirli, Altundag, Yildirim, Tekcan Sanli, & Saatci, 2021; Laurendon et al., 2020; Strauss, Lantos, Heier, Shatzkes, & Phillips, 2020). On the other hand, no significant difference in OB volume between a COVID-19–related anosmia group and a general postviral anosmia group has been reported (Altundag et al., 2020) and a post-mortem tissue examination from patients with severe COVID-19 indeed found virus in the olfactory nerve but only in the leptomeninges layer of the OB (Khan et al., 2021), leaving no consensus of whether COVID-19 is a neurotropic virus.

Problems with most published studies assessing neurological effects of COVID-19 are the focus on single cases or acute, particularly severe cases, where olfactory dysfunction is known to often be less severe (von Bartheld et al., 2020); all without measure of the individual’s state prior to infection and without control group. Without baseline pre-COVID-19 assessment or control group, effects could be population wide or reflect preexisting COVID-19 risk factors.

Recently, a large study on the UK-biobank material provided the first pre-post demonstration of COVID-19 related cerebral alteration in a non-hospitalized sample. By comparing MR assessments of individuals before and after confirmed seropositivity for SARS-CoV-2 to those of seronegative control individuals, loss of gray matter in areas connected to primary olfactory areas, such as the orbitofrontal cortex and insula cortex, were demonstrated (Douaud et al., 2021). Unfortunately, potential insults to the olfactory bulb were not assessed and olfactory function was not measured.

To understand the central mechanisms of COVID-19 related olfactory dysfunction in humans, full psychometric assessment of olfactory function and measures of morphology of the central olfactory system is needed; preferably from the same individual both before and after infection and with the inclusion of a relevant control group for comparison. That said, inducing COVID-19 for experimental demand is ethically questionable, and to our knowledge, no such dataset has been presented.

However, in the months leading up to the first COVID-19 outbreak in Stockholm (late 2019 – early 2020), we acquired full scale psychometric olfactory assessments and structural MR images from a group of healthy individuals as a control group in a study assessing neural effects of olfactory dysfunction. One year into the pandemic and, importantly, before the general vaccination program was initiated in Sweden, we surveyed these individuals about whether they had been infected with the SARS-CoV-2 virus and invited them back for a second assessment of olfactory function and MR scanning. Of those willing to participate in the post study, we recruited 9 individuals who had since the first study suffered from COVID-19, and 12 who had not. In this natural experiment with pre-and post-measures in both COVID-19 affected individuals and a comparable control group, we aimed to determine whether COVID-19 alters olfactory function and the morphology of cerebral areas associated with olfactory processing: the olfactory bulb, anteriorposterior piriform cortex, and central areas of the orbitofrontal cortex. Secondary, we aimed to assess potential links between morphological changes and changes in olfactory functions due to COVID-19. In addition, we measured functional connectivity between olfactory areas in both groups in the post study.

Specifically, we had four hypotheses. We hypothesized that after COVID-19 infection: (1) olfactory bulb volume decreases, (2) gray matter volume in olfactory cortex (anterior and posterior piriform cortex) decreases, (3) functional connectivity during rest between olfactory cortical regions (anterior and posterior piriform cortex and orbitofrontal cortex) is unaffected, and (4) olfactory function is decreased. Critically, we preregistered our hypotheses and analyses prior to assessing the data.

## METHOD

All methods and analyses are according to our preregistration (https://aspredicted.org/wr4d9.pdf) unless otherwise explicitly stated.

### Participants

All participants from a previous study, referred here to as the “Pre-study” (n = 52) that took place September 2019 to February 12th, 2020 (before the COVID-19 outbreak in Stockholm), were inquired about whether they had contracted COVID-19 at some point during the pandemic and whether they consented to participating in a follow-up study (Post-study). In the Post-study, we included both participants who had contracted COVID-19 during the pandemic, and participants who had not. A total of 40 individuals (77%) responded, out of which we classified 9 (6 women) as have been infected with COVID-19 (C19+). Seven out of these reported having tested positive, either for ongoing infection (n = 3) and/or antibodies (n = 5), and one had been diagnosed with COVID-19 without being tested (this happened before testing without hospital admittance was available in Sweden). All 8 agreed to participate in the post study. None had been severely ill or needed hospital care. We also recruited 12 control participants (5 women) who were classified as not having contracted COVID-19 (C19-) based on negative antibody tests. The remaining responding participants from the pre study either did not wish to participate in the post study, were not clearly classifiable as either C19+ or C19-or belonged to the over-represented C19-category. One individual in the C19-group tested positive for ongoing infection a few weeks after the post study. Once recovered, she participated in the Post-study once more; this time as a C19+ participant. This participant thus contributed both to the C19- and C19+ groups, resulting in n = 12 and 9, respectively. The C19+ participants were on average 38 years old (SD = 8, range 30-51) and the C19-participants were on average 33 years old (SD = 7, range 26-49) at the pre study. The post study took place from January to April 2021, between 3 weeks and 12 months after infection for the C19+ group (mean = 7 months; SD = 4). In the Pre-study, all participants reported having normal or corrected-to-normal vision, hearing, and olfactory function.

All procedures were in accordance with the Declaration of Helsinki and approved by the Swedish Ethical Review Authority. All participants provided written informed consent prior to participating in both the Pre- and the Post-study.

### Psychometric odor assessment

Individual olfactory performance was assessed after the MRI data acquisition in both the Pre- and Post-study. We measured odor detection threshold, olfactory quality discrimination, and cued olfactory identification, using the validated Sniffin’ Sticks testing set (Hummel, Sekinger, Wolf, Pauli, & Kobal, 1997) which consists of felt-tipped pens filled with odorants; see details below. The sum of the threshold, discrimination, and identification (TDI) scores was used as an estimate of olfactory function.

#### Odor detection threshold

Absolute sensitivity was assessed for the odor n-Butanol using a three-alternative forced-choice staircase procedure with seven reversals in a 16-step binary dilution series.

#### Odor quality discrimination

Sixteen triplets of pens were presented to the participant. Each triplet consisted of two pens with identical odorants and one with an odorant of different quality. Participants were tasked with identifying which odor in each triplet had a different quality than the other two.

#### Cued odor identification

Olfactory identification performance was assessed with a forced-choice cued identification task using 16 different odorants. Each odor was presented together with a cue card listing four alternative odor labels, and participant picked the label which best described the quality of the presented odor.

The possible range for the threshold measure was 1-16, and for the discrimination and identification measures 0-16, with higher scores indicating better performance, rendering a maximum TDI score of 48. Participants were blindfolded during the threshold and discrimination tests.

### Neuroimaging acquisition and processing

#### MR Acquisition

For both sessions, the same Siemens 3 T Magnetom Prisma MR scanner with a 20-channel head coil was used. We acquired structural images in both studies using identical protocols with a 3D MPRAGE T1-weighted sequence (208 slices, TR = 2300 ms, TE = 2.89 ms, FA = 9°, voxel size = 1 mm × 1 mm × 1 mm, FoV = 256 × 256 voxels). To assess potential effects of COVID-19 on olfactory functional connectivity, the Post-study (but not the Pre-study) included a 12-minute functional restingstate scan using an echo-planar imaging sequence (56 slices, TR = 1700 ms, TE = 30 ms, FA = 70°, voxel size = 2.2 mm × 2.2 mm × 2.2 mm, FoV = 94 × 94 voxels).

#### Image quality assessment

For one participant in the C19+ group, all neuroimaging data were excluded from analysis due to excessive motion artifacts in both the Pre- and Poststructural images which made delineation of the olfactory bulbs problematic.

#### Volumetric measures

Olfactory bulb (OB) volume was assessed manually for each structural image and hemisphere. Data from both sessions for each participant were assigned to one of two experienced neuroradiologist raters (co-authors VL and MB) who were naïve to whether participants belonged to the C19+ or C19-groups.

For the full cerebral cortex, voxel-based morphometry (VBM, J Ashburner & Friston, 2000) analysis was done using the longitudinal pipeline in the Computational Anatomy Toolbox version 12.8 (CAT12; http://www.neuro.uni-jena.de/cat/) for SPM12 (Wellcome Trust Centre for Neuroimaging, UCL; http://www.fil.ion.ucl.ac.uk/spm) in MATLAB 2019b (The MathWorks, Inc., Natick, Massachusetts, USA). Our preregistered analysis plan was based on a cross-sectional publication (Peter et al., 2020), whereas the current study has a longitudinal nature. Consequently, we opted to use the longitudinal pipeline in the CAT12 toolbox, which entails a few minor deviations from the preregistered plan, such as additional intra-subject processing steps and the use of Geodesic Shooting (John Ashburner & Friston, 2011) instead of DARTEL for spatial registration. The recommended default longitudinal VBM processing pipeline of the T1-weighted images in CAT12 was used, including intra-subject realignment and bias field corrections, segmentation into gray matter, white matter, and cerebrospinal fluid, and realignment and normalization into MNI-space using Geodesic shooting with 1.5 mm isotropic voxels, with modulation to preserve gray matter volume.

#### Connectivity measures

Data preprocessing and denoising of functional images was done using SMP12 in Matlab 2019b and the CONN functional connectivity toolbox version 20.b following the steps outlined in (Peter et al., 2021). Preprocessing included slice timing correction, realignment, coregistration of structural and functional data, and normalization to MNI-space. Denoising included removal of five principal components from the white matter and cerebrospinal fluid signals, respectively (Iravani et al., 2021), linear detrending, bandpass filtering (0.01-0.1 Hz), and regression of the six realignment parameters, their first derivatives, and volumes with a framewise displacement > 0.5 mm (Power et al., 2014). No group differences in motion were demonstrated based on Welch’s t-tests of mean framewise displacement (C19+: 0.2 mm; C19-: 0.18 mm; *t*(16.4) = 1.25, *p* = .31) or number of volumes with a framewise displacement > 0.5 mm (C19+: 6.1; C19-: 4.9; *t*(13.7) = 0.39, *p* = .7).

#### Creation of regions of interest

Three regions of interest (ROIs) were included to assess potential COVID-19-related alterations in cortical structure and functional connectivity in areas associated with olfactory processing: the anterior piriform cortex (APC), posterior piriform cortex (PPC), and orbitofrontal cortex (OFC). All ROIs were based on a published olfactory activation likelihood analysis (Seubert, Freiherr, Djordjevic, & Lundström, 2013). To restrict their extension to core processing areas, the piriform ROIs were restricted by an anatomical piriform ROI based on a previously performed manual delineation (Porada, Regenbogen, Seubert, Freiherr, & Lundström, 2019) whereas the OFC ROI was restricted to the frontal pole and orbitofrontal regions of the Harvard-Oxford cortical structural atlas, further described by Seubert et al. (2013). Auditory and visual ROIs corresponding to the functions of the olfactory ROIs were included as control regions for the functional connectivity analysis: The primary auditory cortex (A1), higher order auditory cortex (hAC), primary visual cortex (V1), and the lateral occipital complex (LOC); all defined in Porada et al. (2019).

### Statistical analyses

Change scores were calculated as (post) – (pre) for each individual for the measures that were acquired in both sessions.

#### Olfactory function

The hypothesis of reduced olfactory function in the C19+ group compared to the C19-group was assessed with a one-sided Welch’s t-test on the olfactory TDI change scores. For the subjective change in olfactory function, we had not preregistered any analysis and therefore instead used a two-sided t-test.

#### Volumetric measures

For each structural image, we computed the average OB volume across the left and right hemisphere. Our hypothesis of reduced OB volume in the C19+ compared to the C19-group was assessed using a one-sided Welch’s t-test on the change scores.

For each structural image, gray matter volume in the two preregistered olfactory ROIs (APC and PPC) as well as in an additional olfactory ROI (OFC) was extracted separately for the left and right hemispheres and then averaged over hemispheres. Our hypothesis of reduced gray matter volume in the C19+ group compared to the C19-group was tested using one-sided Welch’s t-tests on the change scores. In addition, an exploratory whole-brain group comparison of gray matter volume was done using a mass-univariate approach of voxel-wise tests of the interaction between the factors Group (C19+/C19-) and Time (pre/post) with a threshold of *p* < .001 and minimum cluster size of 10 voxels.

#### Connectivity measures

The functional imaging for the connectivity analysis was only done in the Post-study. For each participant, we extracted the BOLD time series from each of the three olfactory ROIs as well as from the two auditory and two visual control ROIs. Functional connectivity between the regions was calculated based on pairwise Pearson’s correlation within the three sensory systems separately, i.e., five correlation measures in total. The correlation values were Fisher’s z-transformed for statistical comparisons between the two groups, which was performed using two-sided Welch’s t-tests.

## RESULTS

We first assessed the objective change in olfactory function due to COVID-19 by comparing the change in threshold and TDI scores of the C19+ and C19-groups. We hypothesized that the C19+ group would demonstrate a larger reduction in olfactory performance than the control group, and therefore applied the preregistered one-sided Welch’s t-test to the change scores. However, contrary to our hypothesis, there was no significant difference between the groups in threshold, *t*(12.1) = .97, *p* = .82, hedge’s *g* = .44, or TDI, *t*(9.4) = .60, *p* = .72, hedge’s *g* = .29. Contrary to this lack of apparent objective difference between the C19 + and C19 - groups in olfactory function, 4 out of the 9 participants in the C19+ group did experience subjective olfactory and/or gustatory dysfunction at the time of the post study, including one case of parosmia (raw onion smell was replaced by sweat smell). An additional 2 participants reported that they had experienced problems in the acute phase of infection but had since recovered. The self-estimated overall olfactory function, as compared to around the time of the Pre-study, ranged from 50% to 100% (mean = 87.5%, SD = 17.9) in the C19+ group whereas for the C19-group, 100% of participants rated themselves as experiencing no difference in olfactory performance.

We then determined whether COVID-19 might lead to a long-term reduction in olfactory bulb (OB) volume after COVID-19. On average, the OB volume in the C19+ group was reduced with 14.6% (SD = 26.8%) in the post-COVID measure compared to their pre-COVID measure while in the C19-group, the corresponding number was a 2.3% (SD = 23.2%) volume reduction (Figure 1). Our preregistered one-sided Welch’s *t*-test on the absolute change scores showed that this difference between groups was non-significant according to our significance cut-off alpha value (.05), *t*(15.2) = 1.3, *p* = .1, Hedge’s *g* = .58. However, it is interesting to note that in the C19+ group, 87.5% of participants (7 out of 8) demonstrated a reduction in OB volume (mean= 22.7%, SD = 15.2). Moreover, when assessing the likelihood that the observed reduction in OB volume reduction in the C19+ occurred due to change using a non-preregistered binominal test, we found that the probability that 7 or more C19+ participants would demonstrate a reduction was p = .035, z = 1.76.

**Figure 1.**
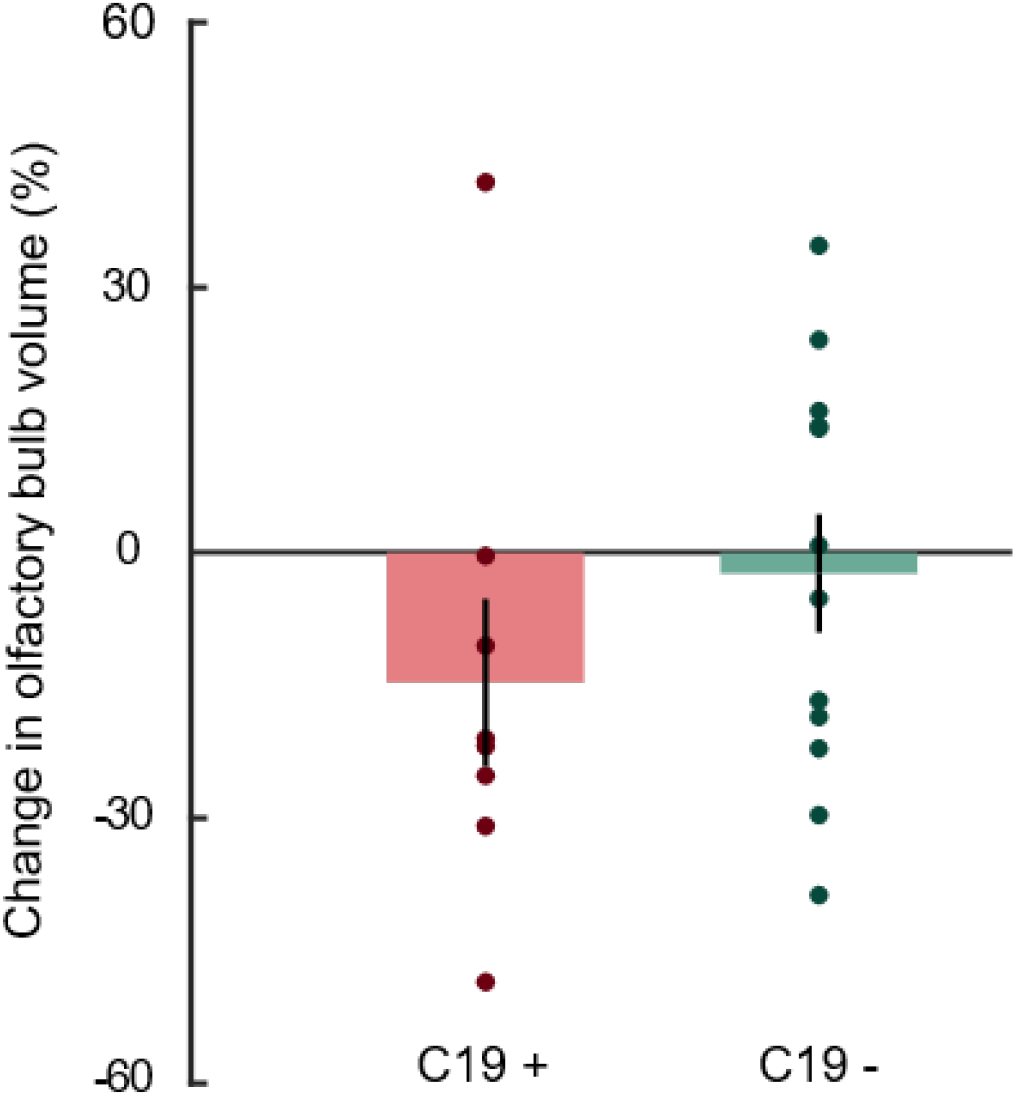
Mean (bars) and individual (dots) percentage of olfactory bulb volume change scores between post-pre COVID-19 for the C19 + and C19-groups. Error bars denote ±1SEM.

Next, we assessed whether COVID-19 leads to loss of gray matter volume in central olfactory areas by determining whether the gray matter volume change-score was different in the C19+ group compared to the C19-group. The nominal values were similar between the two groups in change-score and we did not find any statistical differences in the three ROIs with all p-values above .27 (Figure 2). Finally, in an exploratory and not preregistered analysis, we assessed whether there were signs of volumetric changes to any non-hypothesized brain areas by performing a whole-brain contrast between C19+ and C19-groups. No voxels survived the set statistical threshold of *p* < .001 and cluster size of ≥ 10.

**Figure 2.**
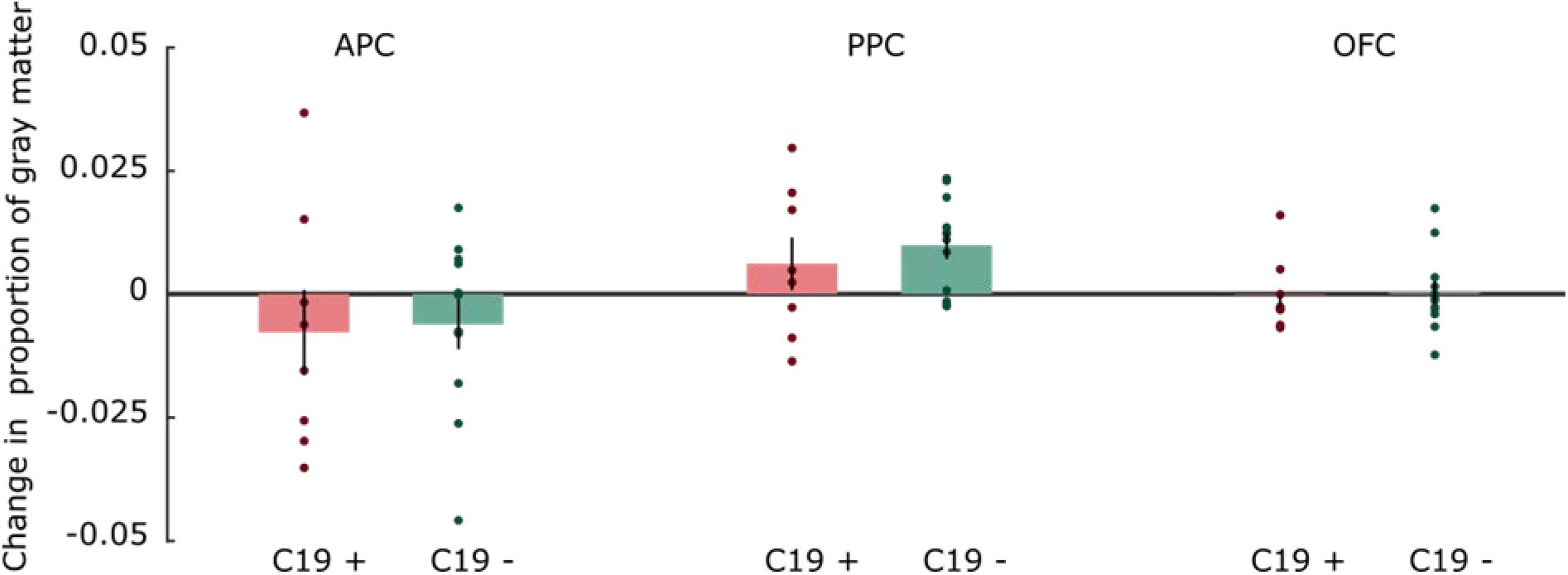
Mean (bars) and individual (dots) pre-post change scores for proportional change in gray matter volume within the C19 + and C19-groups, separately for each ROI. Error bars denote ±1SEM.

Finally, we assessed whether COVID-19 could be linked to alterations in resting-state functional connectivity between core olfactory regions. We found that the C19+ group demonstrated an increase in functional connectivity between the orbitofrontal cortex and the anterior piriform cortex, *t*(14.6) = 3.92, *p* < .005, *g* = 1.73, as well as the orbitofrontal cortex and the posterior piriform cortex, *t*(12.1) = 3.07, *p* < .01, *g* = 1.42 (mean *r* = .26 and *r* = .22, respectively) compared to the C19-group (mean *r* = .12 and *r* = .1, respectively). However, we found no significant group differences in functional connectivity between the closely located anterior and posterior piriform cortex, *t*(13.6) = 0.15, *p* = .89, *g* = .07 (Supplementary Figure S1). Likewise, we found no significant group differences in functional connectivity between our control regions, namely auditory primary and higher order auditory cortex, *t*(12.9) = 1.47, *p* = .17, *g* = .67, and primary visual cortex and lateral occipital complex, *t*(16.4) = 0.65, *p* = .53, *g* = .28.

We did not hypothesize that COVID-19 would produce an increase in functional connectivity between piriform cortex and orbitofrontal. Our preregistered hypothesis was that there would be no difference in functional connectivity between groups and past research in sensory deprivation-related changes in connectivity from basic sensory areas tend to indicate that sensory loss leads to a decrease in connectivity (Yu et al., 2008). However, it could be hypothesized that this increase in connectivity to an area associated with conscious recognition of the odor (Li et al., 2010) might be produced by the increase in effort needed to process odors after COVID-19. We therefore performed an additional and not preregistered analysis where we assessed whether the increased connectivity between the orbitofrontal cortex and the two subdivisions of the piriform cortex was related to the participants’ perceived subjective change in olfactory ability post COVID-19. Spearman’s correlation between perceived olfactory ability and connectivity values between the orbitofrontal and the anterior and posterior piriform cortex, respectively, was calculated for the C19+ group. However, no significant correlation between the perceived olfactory ability and orbitofrontal-anterior piriform connectivity, *r*_S_ = -.39, *p* = .33, or the orbitofrontal-posterior piriform connectivity was demonstrated, *r*_S_ = -.16, *p* = .7.

## DISCUSSION

Facing the current pandemic, worldwide efforts have been made to better understand the links between COVID-19 and olfactory dysfunction. Here, we aimed to take advantage of a unique group of participants that allowed us to assess COVID-19-dependent effects on the morphology of the human olfactory bulb (OB) and cerebral olfactory areas using a within-subject design with a comparable control group.

In line with our pre-registered hypothesis, we observed a consistent decrease of OB volume in 7 out of 8 measured individuals within the C19+ group with an average of about 14% decrease in volume on average of 7 months after their infection by the SARS-CoV-2 virus. In the control group, 6 had an increase and 6 a decrease over time with an av6erage decrease of about 2% in OB volume. Although it should be clearly noted that this was not a statistically significant difference between groups (p = .1) according to the pre-registered alpha cut-off value for our inference analyses, it is interesting to note that a binominal test indicates that the outcome is unlikely to occur due to chance as well as demonstrating a medium effect size, even when including a deviating value with large increase in OB volume. Based on this, it can be speculated whether the lack of clear significant effects according to our preregistered analyses plan is mainly due to the small sample size, regulated by the unique and restricted population, and the fact that one individual in the C19+ group demonstrated a large increase in OB volume. Unfortunately, because olfactory-focused neuroimaging studies where scarce in close proximity to the onset of the pandemic and a large size of the adult Swedish population are at this time vaccinated, it is not possible to increase the sample size. Although multiple past studies have demonstrated that OB volume is modulated by changes in olfactory performance (Negoias, Pietsch, & Hummel, 2017), the mechanism allowing this plasticity is not known. Studies in animal models have demonstrated that neurogenesis can occur in the OB (Bergmann et al., 2012) but studies in human cadavers have not supported this phenomenon. A more straightforward mechanism that might explain the link between the fast changes in OB volume that olfactory training is known to induce is potential changes in the OBs vascularization. Recent data suggest that nearly all individuals infected by the SARS-CoV-2 virus, regardless of whether symptomatic or not, experience endothelial cell death which cause microvascular damage to tissue along the olfactory pathway (Meinhardt et al., 2021; Wenzel et al., 2021). Given the flexibility of the OBs vascularization and close link to amount of olfactory input (Korol & Brunjes, 1992), this suggest that olfactory training might help alleviate OB morphological loss due to COVD-19. However, given the link between olfactory input and OB volume, we cannot dissociate between direct effects from the SARS-CoV-2 virus and that from a potential reduction in olfactory sensory input. That said, although several participants in the C19+ group reported subjective changes in olfactory functions, we did not find any statistically significant changes in objective olfactory performance.

We found no evidence that a COVID-19 infection causes long-term insult to cerebral areas of the olfactory system (the piriform and orbitofrontal cortex) but we did demonstrate a significant increase of functional connectivity between the orbitofrontal cortex and both anterior and posterior piriform cortex. These outcomes were all conflicting with our pre-registered hypothesis. However, absence of clear COVID-19-related morphological changes to piriform cortex, often referred to as primary olfactory cortex, was also demonstrated in the large study on the UK-biobank material where pre- and post-COVID-19 infection data were included. In contrast, the UK-biobank study found COVID-19-related reduction in gray matter within the orbitofrontal cortex where we found no differences (Douaud et al., 2021). The two studies differ in sample size, scanning parameters, and location of our olfactory-related ROIs. That said, although our scanning parameters were optimized for achieving good signal in the orbitofrontal areas of the brain and there were no trends of a difference in our obtained results, larger sample size might have allowed us to replicate the finding of Douaud and colleagues (2021). Nonetheless, our lack of significant results supports the emerging consensus that COVID-19 does not cause long-term morphological alterations to the olfactory cortex of such magnitude that it can be clearly demonstrated on average 7 months after infection. In the majority of studies on COVID-19 influence on the olfactory system, olfactory function has either not been assessed, assessed using subjective self-reports, or assessed with cued olfactory identification performance. Self-reports are a poor measure of olfactory function (Landis, Hummel, Hugentobler, Giger, & Lacroix, 2003). While most people do notice sudden and complete loss of olfactory function, awareness of a partial loss is far lower than comparable perceptual loss in other sensory modalities like audition and vision. Cued identification performance alone is a crude measure of olfactory function given the use of strong odors and its partial reliance on cognitive and language skills (Hedner, Larsson, Arnold, Zucco, & Hummel, 2010; Larsson, Nilsson, Olofsson, & Nordin, 2004). Further, the difficulty level is largely decided by the similarity between the presented odor and the lures on the cue card rather than by the odor itself. Therefore, to reliably estimate olfactory loss it is necessary to probe several aspects of olfactory function (threshold, discrimination, and identification) using objective tests in addition to self-reports. Nearly all of our included C19+ participants had recovered their olfactory functions at the time of post-covid scanning and although there was a statistical trend that the two groups differed in subjective assessment of their olfactory performance, we found no significant difference in objective olfactory performance. Although unlikely, it is therefore possible that the lack of long-term cortical effects might be due to neural recovery. That said, such neural recovery mechanism has yet to be demonstrated in olfactory areas within humans.

Individuals that had undergone COVID-19 demonstrated an increase in functional connectivity between piriform and orbitofrontal areas when compared to controls; an effect that went against our pre-registered hypothesis and that was not directly linked to differences in individuals’ subjective olfactory functions. We based our hypothesis on previous findings that neither long-term, nor short-term, olfactory deprivation results in group differences in functional connectivity between core olfactory regions (Iravani et al., 2021; Peter et al., 2021). A difference between these two past studies and the present study is that nearly all participants in the C19+ had recovered their objective olfactory functions and the increase in functional connectivity might be associated with this recovery. Whether recovery of olfactory functions produce these changes needs to be explored in future studies.

The present study is in many ways unique in that we were provided with the opportunity to assess effects of COVID-19 infection within subjects, with a matching control group, and using a study design designed for assessing potential neural effects of olfactory dysfunction. Without baseline pre-COVID-19 assessment or control group, effects could be population wide or reflect pre-existing COVID-19 risk factors. That said, the study is limited by the restricted sample size and it should be clear that it does not have the same predicted power as a randomized control study where participants are randomly assigned to conditions. It is possible that C19 status was, to some extent, determined by personality traits or other factors. Nonetheless, our baseline measures of the individual’s state prior to infection and, critically, the inclusion of individuals with only mild to medium COVID-19 symptoms is a strength over studies assessing clinical cases with a more severe symptoms where the incident of olfactory dysfunction is known to be much lower (von Bartheld et al., 2020).

Evidence from both animal and human data have demonstrated that a range of DNA and RNA viruses are first detected in the OB during neurotropic infections of the CNS (Durrant et al., 2016). In line with this notion are recent data suggesting that although wide-spread disease-associated microglia signatures are found in COVID-19 infected patients’ cortex, there are no molecular traces of SARS-CoV-2 in the cortex beyond the OB (Yang et al., 2021), a finding supported by the discover of SARS-CoV-2 virus in the olfactory bulb, but not beyond, in an animal model (de Melo et al., 2021). These findings are further in line with past data suggesting that OB interneurons are not affected by neurotropic coronaviruses (Wheeler, Athmer, Meyerholz, & Perlman, 2017) and that the OB might provide virologic control by clearing viruses rapidly after infection (Kalinke, Bechmann, & Detje, 2011). Our results of tentative long-term morphological effects in the OB, but not olfactory cortex, therefore support the notion that the OB functioning as a immunosensory effector organ during neurotropic viral infections (Durrant et al., 2016).

To conclude, we demonstrate tentative evidence that COVID-19 reduces the volume of the OB with an average of 14% but does not affect gray matter volume of the main cerebral olfactory areas. Although 87.5% of our participants demonstrated a reduced OB volume average of 7 months after COVID-19 and binomial testing suggests that the result is not due to change, our findings did not, however, reach a formal statistical significance threshold.

## Data Availability

All data produced are available online at www.osf.io

https://osf.io/9fcr6/

## CONFLICT OF INTEREST

No conflicts of interests declared.

## ACKNOWLEDGEMENT

Funding provided by the Knut and Alice Wallenberg Foundation (KAW 2018.0152) and the Swedish Research Council (2021-06527), awarded to JNL. Data acquisition supported by a grant to the Stockholm University Brain Imaging Centre (SU FV-5.1.2-1035-15).

## SUPPLEMENTARY MATERIAL

**Figure S1.**
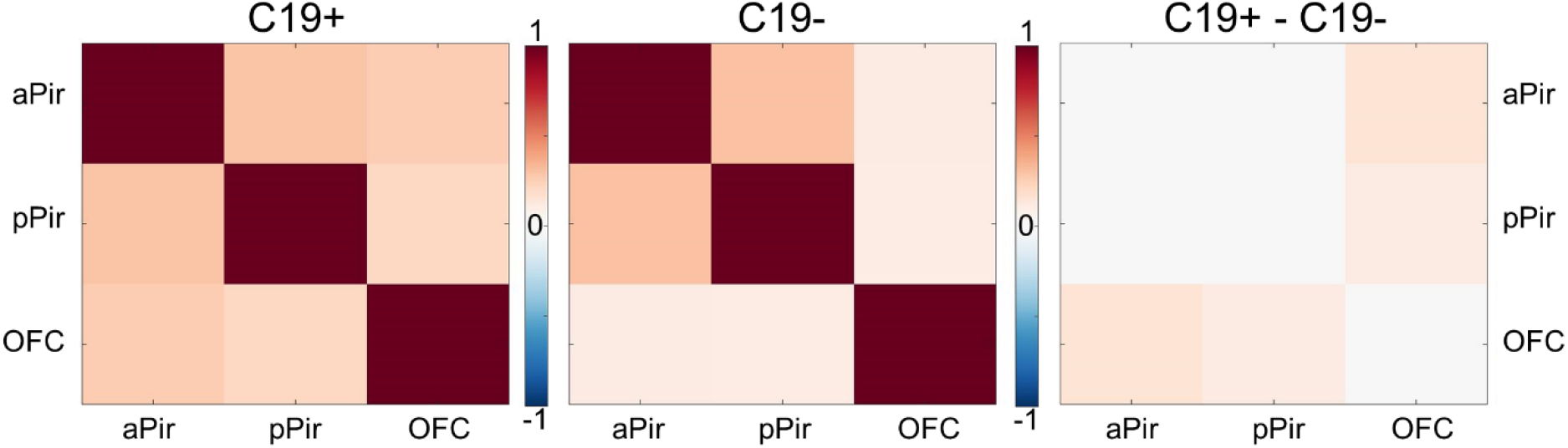
Correlation matrix for the three olfactory ROIs for the C19+ and C19-groups as well as difference between the two groups.

## Notes

### Competing Interest Statement

The authors have declared no competing interest.

### Author Declarations

The Swedish Ethical Review Authority.

